# Active amyloid beta immunization ameliorates synapse loss and phosphorylated-tau accumulation around remaining plaques for up to 14 years after treatment

**DOI:** 10.64898/2026.08.10.26359862

**Authors:** Elizabeth Simzer, Robert I McGeachan, Jane Tulloch, Makis Tzioras, Colin Smith, James AR Nicoll, Delphine Boche, Tara L Spires-Jones

## Abstract

Amyloid plaques, one of the defining features of Alzheimer’s disease, are associated with synapse loss and accumulation of pathological tau in dystrophic neurites and reactive glia in their immediate vicinity. Anti-amyloid-β immunotherapies have been shown to effectively remove a large proportion of plaques from the brain, but whether immunotherapies reduce pathological changes around remaining plaques or plaques that emerge after treatment remains unknown. We examined amyloid plaques, synapses, astrocytes, and phosphorylated tau in *post-mortem* brain tissue from people with Alzheimer’s disease who received Amyloid-β42 immunization in the AN1792 trial (Elan Pharmaceuticals), non-immunized or placebo-treated people with Alzheimer’s disease, and neurologically healthy controls. In non-vaccinated individuals, we observe plaque-associated synapse loss, phospho-tau accumulation, and astrogliosis as previously reported. People who received Amyloid-β_42_ vaccination had reduced pathology up to 14 years after receiving the vaccine including ameliorated plaque-associated synapse loss, less accumulation of phospho-tau around plaques (AT8 and pTau217), lower levels of astrogliosis, and lower levels of phospho-tau associated with synapses. These data indicate that anti-amyloid active vaccines may have lasting beneficial effects even around remaining plaques or plaques formed after immunization.

## Introduction

Amyloid-β (Aβ) accumulation in plaques is one of the defining pathologies of Alzheimer’s disease with plaque accumulation occurring early in disease pathogenesis. Gene variants that cause autosomal dominant, familial Alzheimer’s disease all impact Aβ production or aggregation, leading to the amyloid cascade hypothesis that amyloid initiates disease and decades of therapeutic development aimed at removing amyloid pathology from the brain ^1,2^. Two anti-amyloid immunotherapies, lecanemab and donanemab, have succeeded at modestly slowing cognitive decline in phase III clinical trials and have become available for use to treat Alzheimer’s disease in several countries around the world ^3,4^. While animal models and clinical trials support clearance of amyloid plaques with immunotherapy, plaque load does not strongly correlate with cognitive decline, and the mechanisms underlying cognitive benefit remain incompletely understood.

Plaques are typically surrounded by a halo of toxic oligomeric Aβ, reactive astrocytes and microglia, and local neuron damage including formation of phospho-tau containing dystrophic neurites and synapse loss ^5,6^. Tau pathology burden correlates more strongly with cognitive impairment than amyloid pathology, indicating that resolution of plaque-associated damage including phosphorylated (phospho)-tau containing neurites could contribute to benefits of immunotherapy.

Neuropathological studies after clinical trials of both active and passive immunotherapy to remove Aβ demonstrate substantial removal of amyloid plaque pathology and a reduction of phospho-tau in neurites but not neuronal cell bodies containing tangles ^7–9^. There have not previously been studies specifically examining plaque-associated phospho-tau pathology after Aβimmunotherapy, which is important in understanding why phospho-tau fluid biomarkers and PET signals are lowered by amyloid removal in trials ^4,10–12^.

Synapse loss is the strongest pathological correlate of cognitive decline in Alzheimer’s disease ^13–15^, and our group has demonstrated that the most substantial synapse loss occurs in the direct vicinity of amyloid plaques and is associated with accumulation of oligomeric Aβ within individual synapses ^16–19^. We recently observed that lecanemab positive Aβ is present in synapses around plaques in Alzheimer’s brain tissue, hinting that lecanemab immunotherapy may be able to remove synaptotoxic Aβ ^20^. Plaque-associated synapse loss is thought to be mediated at least in part by glial engulfment of synapses with our data indicating that astrocytes and microglia contain increased levels of synaptic protein around plaques in human *post-mortem* tissue from people with Alzheimer’s disease ^16^.

Due to their recent development, there are not yet studies of brain tissue from people who were treated with lecanemab or donanemab with long-term follow up after treatment which can address how immunotherapy affects plaque-associated pathology. The AN1792 trial, the first active Aβ immunization, provides a unique opportunity to investigate the long-term effects of immunotherapy on plaque-associated pathology. Previous work has shown that AN1792 vaccination was associated with plaque clearance in 14/16 treated Alzheimer’s patients up to 14 years post-vaccination ^8^. In regions with plaque clearance, the burden of tau pathology was also lowered, although overall distribution of tangles was severe in all cases ^8^. It remains unknown whether immunotherapy can reduce pathological changes around remaining plaques or plaques that emerge after treatment. To address this question, we used high-resolution imaging to investigate plaque-associated synapse loss, phospho-tau burden, and astrocytes in brain tissue from the AN1792 immunised cohort and matched placebo or unvaccinated Alzheimer’s cases and controls.

## Methods

### Subjects

Use of human tissue for *post-mortem* studies has been reviewed and approved by the Edinburgh Brain Bank ethics committee and the Academic and Clinical Central Office for Research and Development Medical Research Ethics Committee, a joint office of the University of Edinburgh and NHS Lothian (approval 15-HV-016). The Edinburgh Brain Bank is a Medical Research Council funded facility with research ethics committee approval (11/ES/0022). Ethical approval for the use of the immunised AD cases has been granted by BRAIN UK (REC ref: 19/SC/0217). Table 1 shows summary demographic data of tissue donors. Inclusion and exclusion criteria for each group was as follows: Controls - no neurological or psychiatric clinical diagnoses and limited neuropathological changes at autopsy (tau Braak 0-II), Early Alzheimer’s disease related changes (tau Braak III-IV with amyloid pathology present and limited non-AD pathologies); Late Alzheimer’s - clinical dementia diagnosis and confirmed neuropathological diagnosis of AD with Braak stage V or VI, limited non-AD pathologies See extended data table 1 for case details. UK Brain Bank ID numbers (BBNs) are included for relevant cases in line with our ethical approval which requires publishing BBNs. Participants in the AN1792 trial (NCT00021723) were recruited and treated as previously published.^21^

**Table 1:** Demographic data of brain tissue donors. SD – standard deviation, PMI – *post-mortem* interval, AD – Alzheimer’s disease.

|  | Control<br>(n=13) | AD AN1792<br>(n=11) | AD<br>untreated/placebo<br>(n=16) | Group Comparison |
| --- | --- | --- | --- | --- |
| Sex M (n, %) | 10 (76.9%) | 7 (63.6%) | 9 (56.3%) | X-squared = 1.36, df = 2,<br>p-value = 0.51 |
| Sex F (n, %) | 3 (23.1%) | 4 (36.4%) | 7 (43.8%) |  |
| Age mean (SD) | 80.5 (2.57) | 80.8 (7.88) | 80.6 (9.87) | ANOVA F[2,37]=0.004,<br>p=0.99 |

**Extended data table 1:**
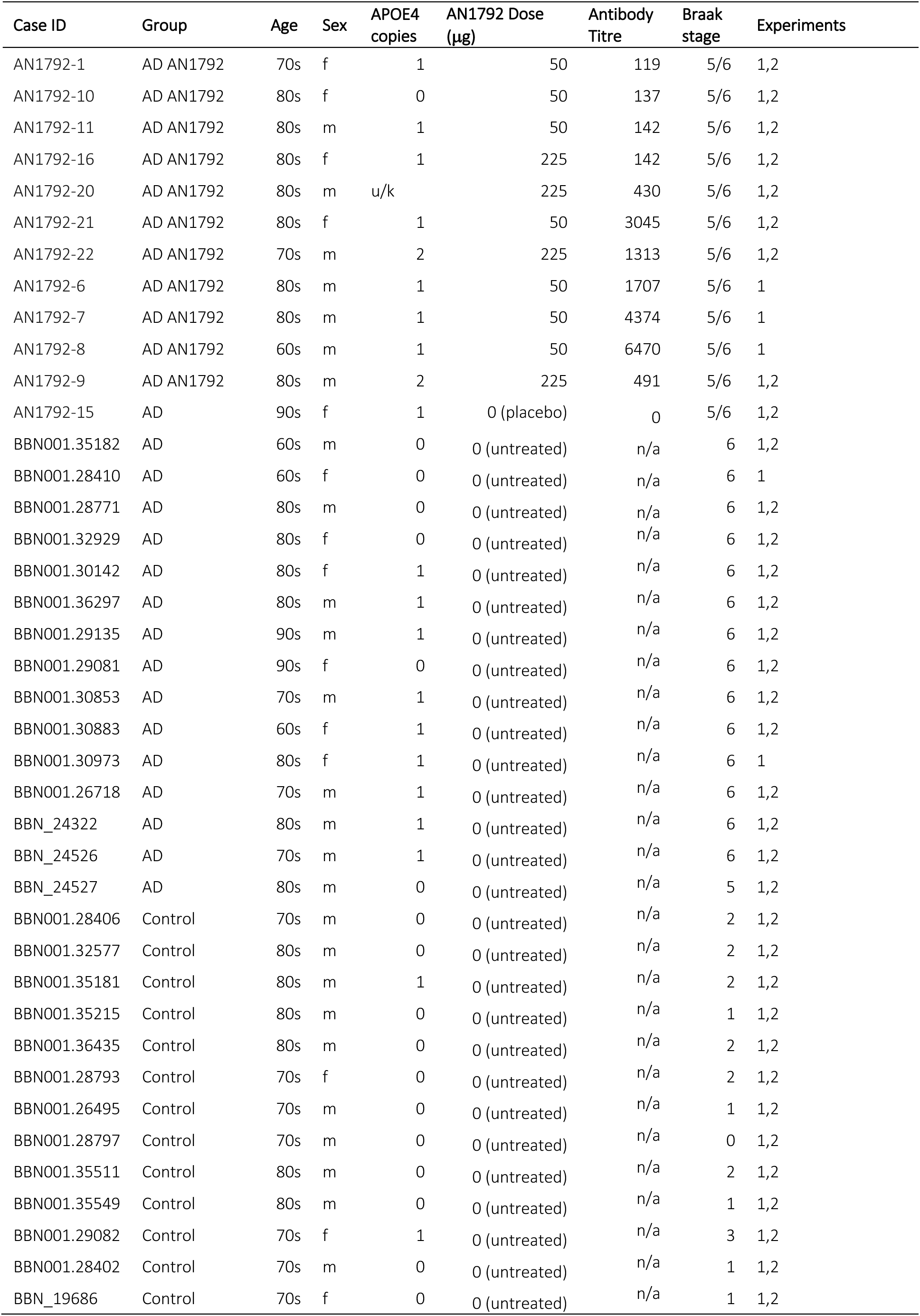
Details of all tissue donors in the study. u/k = unknown, n/a = not applicable, Experiments (1 - Homer, methoxy, pTau217; 2 - Synaptophysin, AT8, GFAP, methoxy)

| Case ID | Group | Age | Sex | APOE4<br>copies | AN1792 Dose<br>(µg) | Antibody<br>Titre | Braak<br>stage | Experiments |
| --- | --- | --- | --- | --- | --- | --- | --- | --- |
| AN1792-1 | AD AN1792 | 70s | f | 1 | 50 | 119 | 5/6 | 1,2 |
| AN1792-10 | AD AN1792 | 80s | f | 0 | 50 | 137 | 5/6 | 1,2 |
| AN1792-11 | AD AN1792 | 80s | m | 1 | 50 | 142 | 5/6 | 1,2 |
| AN1792-16 | AD AN1792 | 80s | f | 1 | 225 | 142 | 5/6 | 1,2 |
| AN1792-20 | AD AN1792 | 80s | m | u/k | 225 | 430 | 5/6 | 1,2 |
| AN1792-21 | AD AN1792 | 80s | f | 1 | 50 | 3045 | 5/6 | 1,2 |
| AN1792-22 | AD AN1792 | 70s | m | 2 | 225 | 1313 | 5/6 | 1,2 |
| AN1792-6 | AD AN1792 | 80s | m | 1 | 50 | 1707 | 5/6 | 1 |
| AN1792-7 | AD AN1792 | 80s | m | 1 | 50 | 4374 | 5/6 | 1 |
| AN1792-8 | AD AN1792 | 60s | m | 1 | 50 | 6470 | 5/6 | 1 |
| AN1792-9 | AD AN1792 | 80s | m | 2 | 225 | 491 | 5/6 | 1,2 |
| AN1792-15 | AD | 90s | f | 1 | 0 (placebo) | 0 | 5/6 | 1,2 |
| BBN001.35182 | AD | 60s | m | 0 | 0 (untreated) | n/a | 6 | 1,2 |
| BBN001.28410 | AD | 60s | f | 0 | 0 (untreated) | n/a | 6 | 1 |
| BBN001.28771 | AD | 80s | m | 0 | 0 (untreated) | n/a | 6 | 1,2 |
| BBN001.32929 | AD | 80s | f | 0 | 0 (untreated) | n/a | 6 | 1,2 |
| BBN001.30142 | AD | 80s | f | 1 | 0 (untreated) | n/a | 6 | 1,2 |
| BBN001.36297 | AD | 80s | m | 1 | 0 (untreated) | n/a | 6 | 1,2 |
| BBN001.29135 | AD | 90s | m | 1 | 0 (untreated) | n/a | 6 | 1,2 |
| BBN001.29081 | AD | 90s | f | 0 | 0 (untreated) | n/a | 6 | 1,2 |
| BBN001.30853 | AD | 70s | m | 1 | 0 (untreated) | n/a | 6 | 1,2 |
| BBN001.30883 | AD | 60s | f | 1 | 0 (untreated) | n/a | 6 | 1,2 |
| BBN001.30973 | AD | 80s | f | 1 | 0 (untreated) | n/a | 6 | 1 |
| BBN001.26718 | AD | 70s | m | 1 | 0 (untreated) | n/a | 6 | 1,2 |
| BBN_24322 | AD | 80s | m | 1 | 0 (untreated) | n/a | 6 | 1,2 |
| BBN_24526 | AD | 70s | m | 1 | 0 (untreated) | n/a | 6 | 1,2 |
| BBN_24527 | AD | 80s | m | 0 | 0 (untreated) | n/a | 5 | 1,2 |
| BBN001.28406 | Control | 70s | m | 0 | 0 (untreated) | n/a | 2 | 1,2 |
| BBN001.32577 | Control | 80s | m | 0 | 0 (untreated) | n/a | 2 | 1,2 |
| BBN001.35181 | Control | 80s | m | 1 | 0 (untreated) | n/a | 2 | 1,2 |
| BBN001.35215 | Control | 80s | m | 0 | 0 (untreated) | n/a | 1 | 1,2 |
| BBN001.36435 | Control | 80s | m | 0 | 0 (untreated) | n/a | 2 | 1,2 |
| BBN001.28793 | Control | 70s | f | 0 | 0 (untreated) | n/a | 2 | 1,2 |
| BBN001.26495 | Control | 70s | m | 0 | 0 (untreated) | n/a | 1 | 1,2 |
| BBN001.28797 | Control | 70s | m | 0 | 0 (untreated) | n/a | 0 | 1,2 |
| BBN001.35511 | Control | 80s | m | 0 | 0 (untreated) | n/a | 2 | 1,2 |
| BBN001.35549 | Control | 80s | m | 0 | 0 (untreated) | n/a | 1 | 1,2 |
| BBN001.29082 | Control | 70s | f | 1 | 0 (untreated) | n/a | 3 | 1,2 |
| BBN001.28402 | Control | 70s | m | 0 | 0 (untreated) | n/a | 1 | 1,2 |
| BBN_19686 | Control | 70s | f | 0 | 0 (untreated) | n/a | 1 | 1,2 |

### Immunohistochemistry staining and imaging

Formalin-fixed paraffin-embedded tissue sections from the parietal cortex were selected from AN1792-treated cases with known plaque clearance ^22^ alongside the same region from age and sex matched control and untreated AD cases. Staining was conducted in two batches with each batch containing cases from both sexes and all diagnostic/treatment groups.

Paraffin-embedded tissue sections were de-paraffinised in xylene (6 min), rehydrated through graded ethanol (100%, 90%, 70%, and 50%; 3 min each), and rinsed in distilled water. Antigen retrieval was performed in 1X citrate buffer (pH 6.0) using a pressure cooker for 3 minutes, then cooled in distilled water. Autofluorescence was reduced by sequential incubation in 70% ethanol, Autofluorescence Eliminator Reagent (Millipore, 2160), 70% ethanol, and 50% ethanol (5 min each), followed by rinsing in distilled water. Tissue sections were outlined with a hydrophobic barrier pen and maintained in 1X PBS throughout the staining procedure. Sections were blocked for 1 hour at room temperature in 1X PBS containing 0.3% Triton X-100 and 10% normal donkey serum, then incubated overnight at 4°C with rabbit anti-synaptophysin (1:200), chicken anti-GFAP (1:750), and mouse anti-phosphorylated tau (AT8; 1:500) or chicken anti-Homer1 (1:500) and rabbit anti-pTau217 (1:1000) diluted in blocking solution.

Following overnight incubation with primary antibodies, sections were washed in PBS and incubated for 1 h at room temperature with Alexa Fluor 488 donkey anti-rabbit, Alexa Fluor 594 goat anti-chicken, and Alexa Fluor 647 donkey anti-mouse or Alexa Fluor 488 goat anti-chicken and Alexa Fluor 594 donkey anti-rabbit secondary antibodies (all 1:500). Antibody details can be found in **Extended data Table 2.** Sections were then incubated for 30 min at room temperature with Methoxy-XO4 (50 μM in PBS containing Kolliphor EL), prepared from a 10 mg mL⁻¹ DMSO stock solution. Residual erythrocyte autofluorescence was quenched with RBC quenching reagent (100 μL per slide, 5 min) (Vector, SP-8400-15), and sections were mounted with ImmunoMount, coverslipped, and sealed with clear nail varnish prior to imaging.

**Extended data Table 2.** Antibody information.

| Primary Antibody Information |  |  |  |  |  |
| --- | --- | --- | --- | --- | --- |
| Name | Host Species | Working Dilution | Supplier | # Cat. ID | Lot ID |
| AT8 | Mouse | 1/500 | Invitrogen | MN1020 | XE3565541 |
| Synaptophysin | Rabbit | 1/200 | Abcam | ab32127 | 1090949-49 |
| GFAP | Chicken | 1/750 | Abcam | ab4674 | 1088394-2 |
| Homer1 | Chicken | 1/500 | Synaptic Systems | 160 006 | 4-29 |
| pTau217 | Rabbit | 1/1000 | Cell Signalling | 51625S | Lot: 1 |
| Secondary Antibody Information |  |  |  |  |  |
| Name | Host Species | Working Dilution | Supplier | # Cat. ID | Lot ID |
| Alexa Fluor 488 donkey anti-rabbit | Donkey | 1/500 | Invitrogen | A21206 | 3148296 |
| Alexa Fluor 594 goat anti-chicken | Donkey | 1/500 | Invitrogen | A11042 | 2720367 |
| Alexa Fluor 647 donkey anti-mouse | Donkey | 1/500 | Invitrogen | A32787 | XH357261 |
| Alexa Fluor 488 goat anti-rabbit | Goat | 1/500 | Invitrogen | A11039 | 2180688 |
| Alexa Fluor 594 donkey anti-rabbit | Donkey | 1/500 | Invitrogen | A21207 | 2747441 |

### Confocal imaging

Image stacks were acquired on a Leica TCS 8 confocal with a 63x oil immersion objective. Image stacks of 15 sections were acquired with 1024×1024 pixel resolution with a z-step of 0.3 μm resulting in stacks of 145 μm x 145 μm x 4.5 μm. Stacks were acquired in regions containing plaques and paired plaque-free regions within the same cortical layer approximately 2 image fields distal to the plaque. Methoxy-XO4 binds beta sheet containing fibrils in pathological protein lesions including staining amyloid plaques and neurofibrillary tangles. Plaques were identified by their distinctive morphology by experienced observers and are easily distinguished from neurofibrillary tangle and neuropil thread staining by their larger size and extracellular roughly circular morphology in classic dense and diffuse patterns. For each case, 10 plaque and 10 paired plaque-free stacks were acquired of regions from all cortical layers in each case where possible. Laser and detector settings were kept identical within paired plaque and non-plaque sites but were changed between regions and cases depending on staining quality.

### Image analysis

In-house custom MATLAB and ImageJ macros were used for all image processing and analyses (available: https://github.com/arraytomographyusers/Array_tomography_analysis_tool). Image stacks were blinded and each channel segmented with either auto local thresholding algorithms or fixed value thresholds. Segmented image stacks containing plaques were used to generate a mask of the largest plaque in the field which was expanded to include a halo of 5 μm from the plaque edge for a “plaque+halo”. The % volume of synapse, phospho-tau, GFAP, and colocalized staining between these segmented channels were measured in plaque-free regions, plaque+halo regions and areas outside the plaque+halo in plaque-containing image stacks.

### Statistical analyses

Demographic data between groups was compared using a Chi squared test for sex and one-way ANOVA for age. Group comparisons of immunohistochemistry data were conducted using linear mixed-effects models, with cohort and sex included as fixed effects. Case was included as a random effect to account for multiple measurements per individual, as previously described. When model assumptions were not met, data were transformed via Tukey’s Ladder of Power. Analysis of variance (ANOVA) with Satterthwaite correction was applied to the models, followed by post hoc pairwise comparisons with Tukey adjustment. All statistical analyses were performed using R (version 4.5.2) using RStudio.

## Results

Sections of parietal cortex (angular gyrus) from people with Alzheimer’s disease who were treated with AN1792 (n=11), people with Alzheimer’s disease who were treated with placebo (n=1) or no treatment (n=15), and age and sex matched controls who did not have neurological disease (n=13) were stained for pre-synaptic terminals (synaptophysin), astrocytes (GFAP) and amyloid fibrils (methoxy X-O4, **Figure 1**). Although controls and AN1792 treated cases had fewer plaques as expected, we focused on areas containing plaques and adjacent plaque-free areas to examine whether plaque-associated pathology is ameliorated by vaccination. As previously reported, some plaques in AN1792 treated cases had a “moth-eaten” appearance indicative of partial clearance (**Figure 1**).

**Figure 1:**
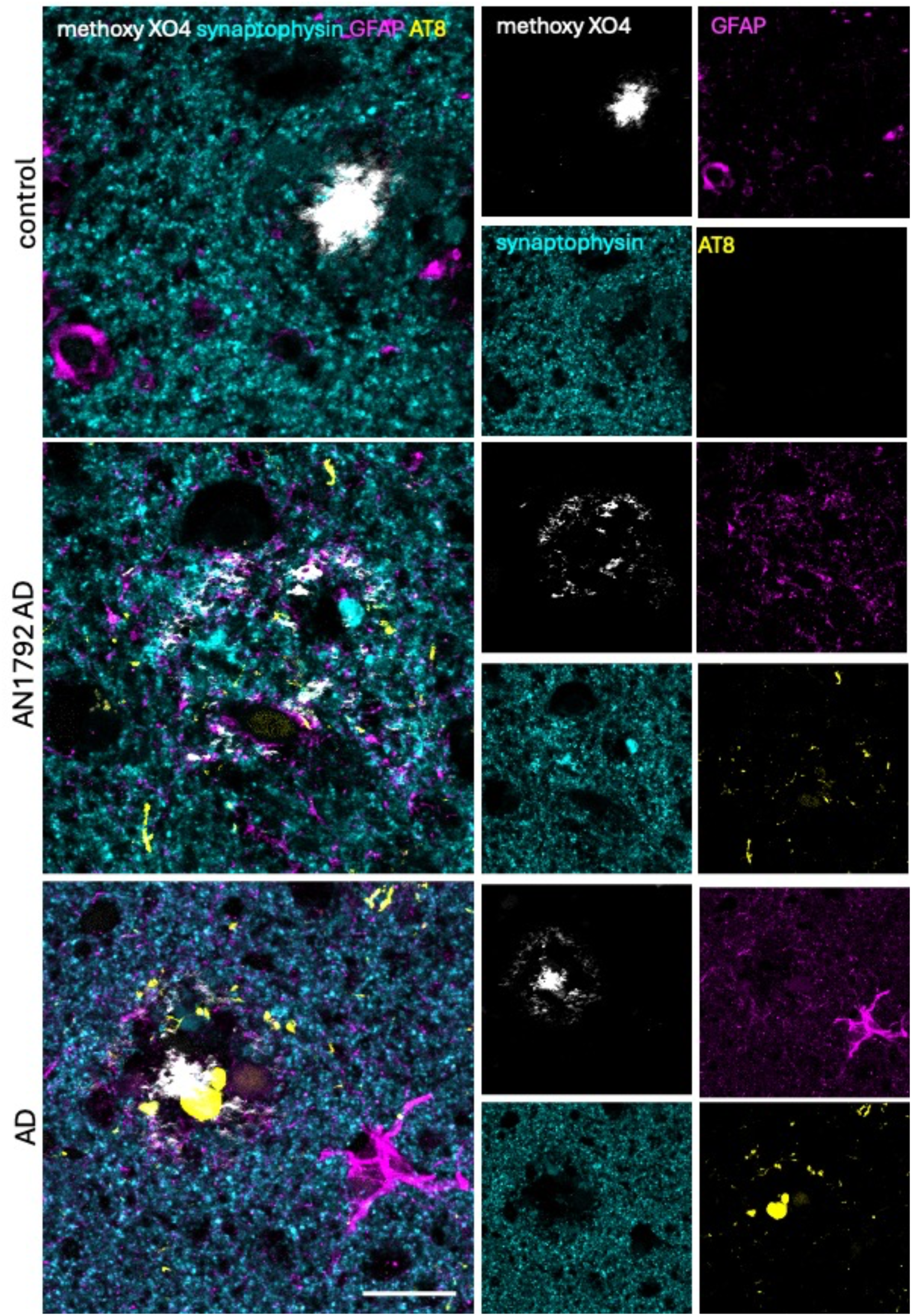
Immunohistochemistry for methoxy XO4 to label plaque figures (grey), synaptophysin (cyan), GFAP (magenta) and AT8 (yellow) was carried out on sections from controls, people with Alzheimer’s disease vaccinated with AN1792(AN1792 AD) and people with Alzheimer’s disease who were not vaccinated (AD). Images are average intensity projections of 15 sections from confocal image stacks with brightness adjusted for clarity. Large images show merged channels with smaller images on right showing each individual channel separately. Note in AN1792 treated cases, some plaques had an “eaten” appearance as in the example shown. Scale bar represents 20 μm.

Plaque-associated synapse loss was assessed by examining the % volume of the tissue occupied by synaptophysin staining in three regions: areas containing plaque staining and the 5 μm of parenchyma around the plaque, areas further than 5 μm from the plaque edge, and the area approximately one field of view (145 μm) distant from the same plaque within the same cortical layer. We observed significant loss of synaptic staining in the immediate vicinity of plaques in untreated or placebo treated Alzheimer’s disease cases and controls, but no loss of synapses around plaques in AN1792 treated tissue donors **(Figure 2)**.

**Figure 2:**
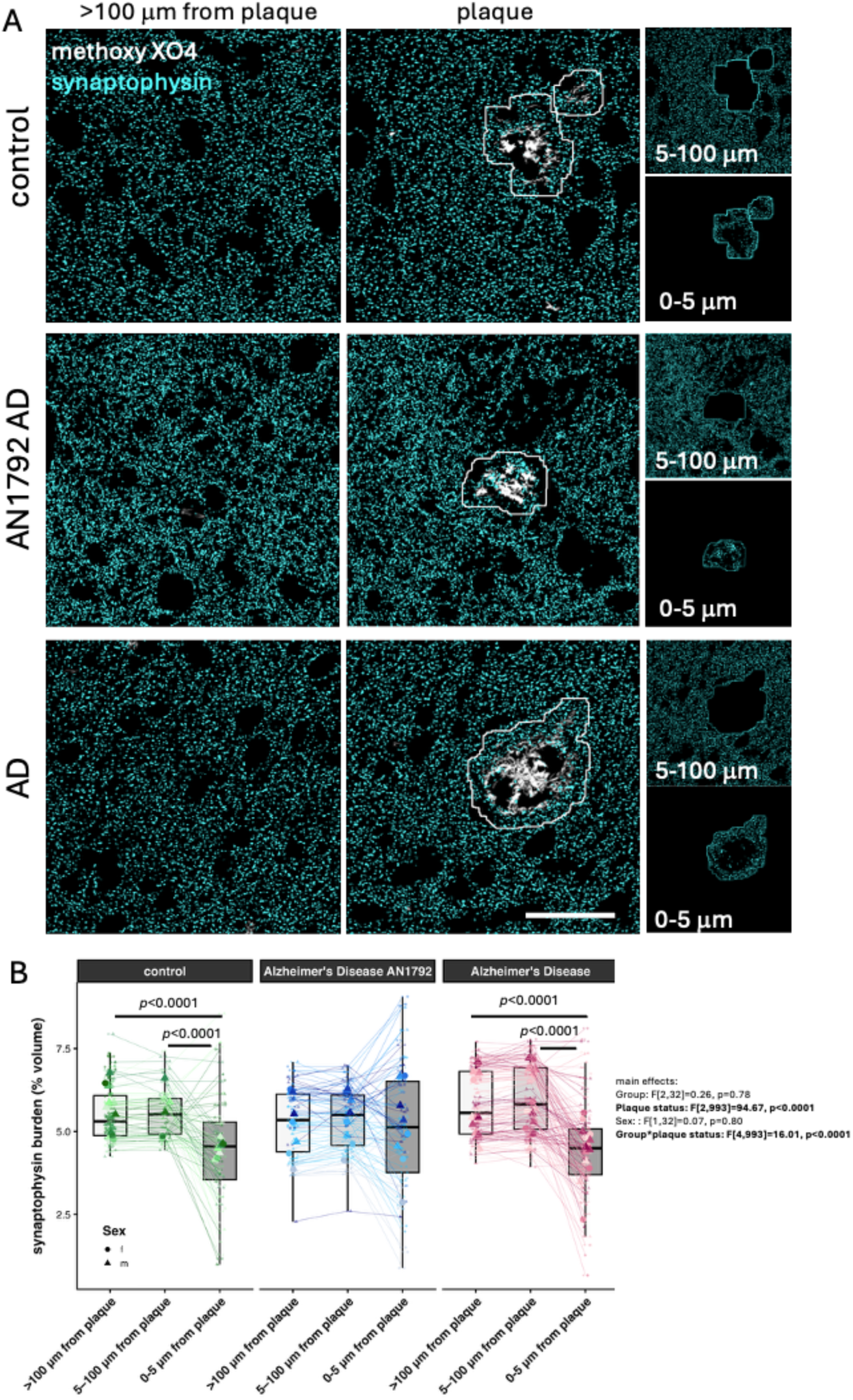
Plaque-associated synapse loss is ameliorated by AN1792 treatment. Confocal image stacks were taken in regions with plaques (methoxy-XO4, white) and in adjacent areas in the same cortical layer at least 100 μm from the plaque (**A**). Image stacks were segmented and the burden of synaptophysin (cyan) staining calculated in the entire field in images far from plaques and in a region of interest containing the plaque and a 5 μm halo around the plaque edge and in the non-plaque/halo region in plaque-containing images (A, right). Quantification and analysis with a linear mixed effects model (group * plaque status + Sex + (1|Case/Image)) reveals plaque-associated synapse loss in controls and subjects with AD but not in people treated with AN1792 (**B**). Images are average projections of 15 sections of segmented image stacks. Scale bar represents 50 μm. Boxplots show medians of individual image pairs as small points connected by lines and case medians as large points. Post-hoc pairwise Tukey corrected p values are shown for groups with significant differences.

We then examined whether phospho-tau (labelled with AT8 antibody), GFAP accumulation, or their colocalization with synaptophysin was affected by AN1792 treatment (**Figure 3**). AT8 burden (% volume) across all regions (plaque and non-plaque) was significantly higher in Alzheimer’s disease than AN1792 treated Alzheimer’s disease and both Alzheimer’s groups had higher AT8 burden than control cases (**Figure 4A**). The area within 5 μm of plaques had the highest level of AT8 staining in Alzheimer’s disease, but this plaque-associated increase was not present with AN1792 treatment. The colocalization of AT8 with synaptophysin staining, indicating likely synaptic accumulation of phospho-tau, was increased in Alzheimer’s disease compared to control and was significantly lower with AN1792 treatment than in the untreated Alzheimer’s group. In the untreated Alzheimer’s group, synaptic tau colocalisation was increased around plaques with the highest levels in the region 5-100 μm away from the plaque edge. Plaque-associated synaptic tau colocalisation was almost absent in the control group and significantly decreased near plaques in the AN1792 treated group (**Figure 4B**). Astrocyte burden was increased in Alzheimer’s disease compared to the control group and was significantly lower in AN1792 treated compared to untreated groups (**Figure 4C**). There was a significant effect of plaque distance across all groups in GFAP burden. Colocalization between AT8 and GFAP indicating astrocyte engulfment of phospho-tau was increased in the Alzheimer’s disease group compared to the control group with the highest levels 5-100 μm from plaques (**Figure 4D**). There were no differences between control and AN1792 treated Alzheimer’s groups in GFAP-AT8 colocalization. There were no significant differences between groups in GFAP colocalization with synaptophysin (data not shown).

**Figure 3:**
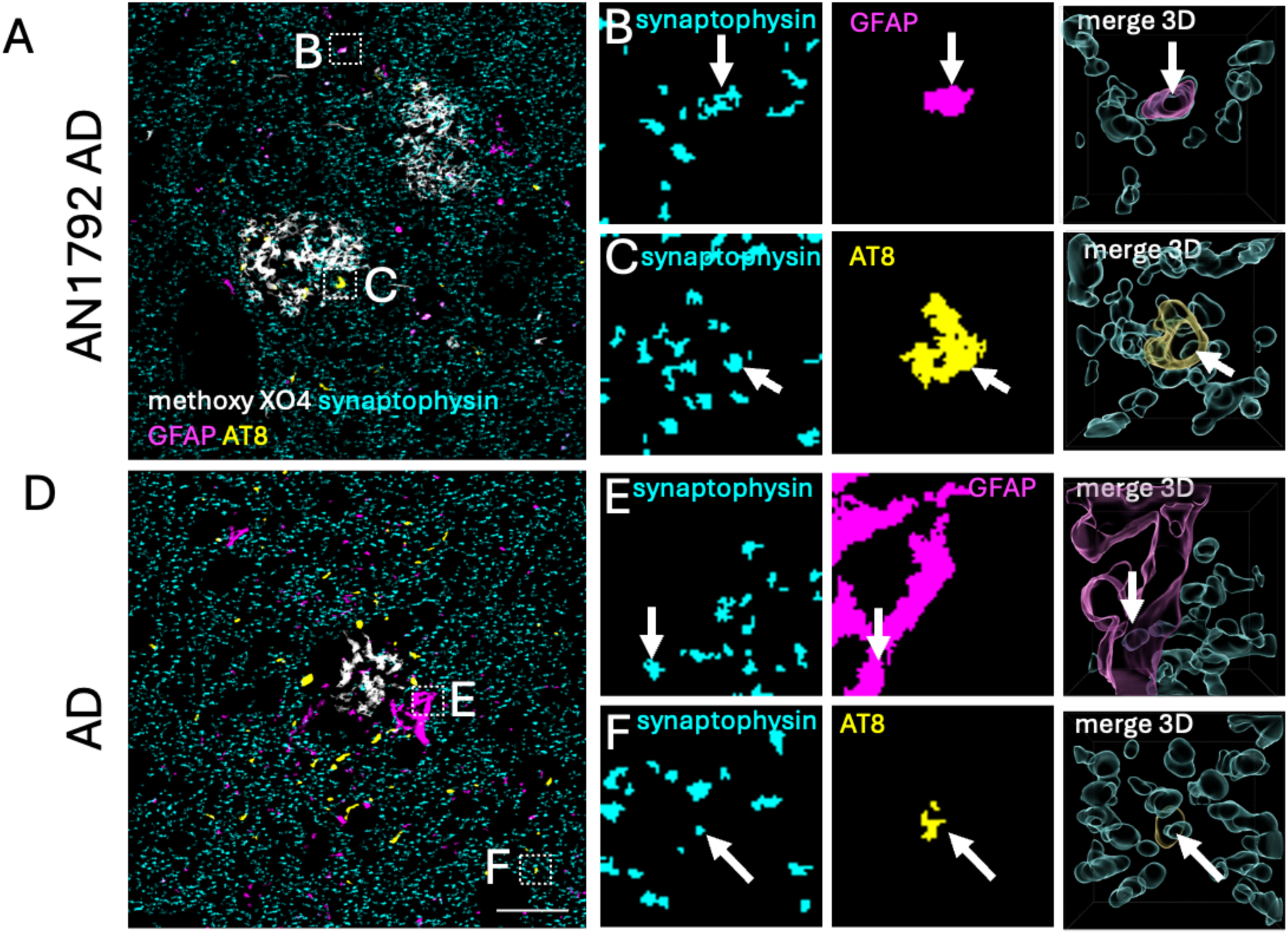
Plaque-associated tau pathology and astrogliosis. Confocal image stacks from AN1792 treated (A-C) unvaccinated people with AD (D-F) were analysed for accumulation of AT8 positive phospho-tau (yellow), GFAP (magenta), and synaptophysin staining (cyan) relative to distance from methoxy-XO4 positive plaques (white). Large images are average projections of 15 sections of segmented image stacks, insets show single section images of each channel and a 3D reconstruction of all merged channels to demonstrate colocalization. Scale bar represents 20 μm insets are 10 x 10 μm.

**Figure 4:**
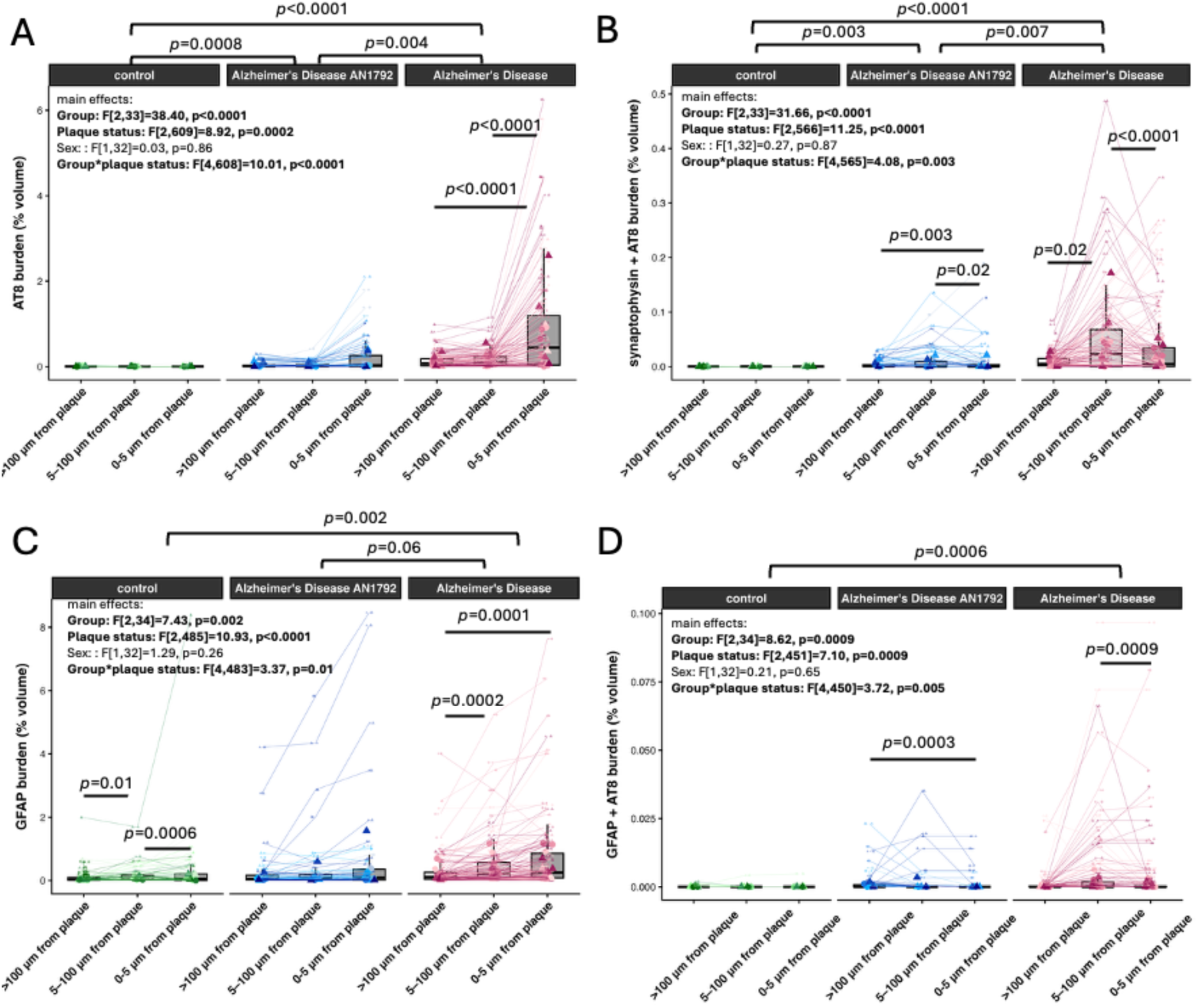
Plaque-associated tau pathology and astrogliosis are reduced by AN1792 treatment. Quantification and analysis with a linear mixed effects model data (group * plaque status + Sex + (1|Case/Image)) reveals plaque-associated AT8 accumulation in AD but not in people with AD vaccinated with AN1792 or controls (**A**). Synaptic phospho-tau accumulation, measured as the burden of colocalized AT8 and synaptophysin was lower with AN1792 treatment (**B**). GFAP burden was higher in people with Alzheimer’s disease than in controls and there was a trend towards reduction of GFAP burden with AN1792 treatment (**C**). Colocalisation of AT8 positive tau with astrocytes was significantly higher in AD compared to control individuals with no difference between AN1792 treated AD cases and either controls or AD individuals (**D**). Boxplots show medians of individual image pairs as small points connected by lines and case medians as large points. Post-hoc pairwise Tukey corrected p values are shown for groups with significant differences. All statistical models were run on Tukey transformed data.

We examined excitatory post-synapses (stained with homer 1 antibody) and phospho-tau 217 accumulation around plaques (**Figure 5**). Similar to the presynaptic marker, we observed synapse loss around plaques in both untreated Alzheimer’s disease and control cases. In AN1792 cases, there was a slight but significant *increase* post-synaptic staining in the immediate vicinity of plaques, possibly reflecting synaptic regrowth or compensation (**Figure 6**). pTau217 staining burden was increased in people with Alzheimer’s disease compared to controls and reduced by AN1792 treatment compared to the Alzheimer’s group although the treated group remained higher than controls. In both AN1792 treated and untreated Alzheimer’s disease, pTau217 staining was increased around plaques. AN1792 treatment was associated with reduced colocalization of pTau217 with post-synapses compared to the Alzheimer’s disease group although still higher than controls. There was very little colocalization between pTau217 and homer in controls (median 0 at all plaque distances). With respect to plaque proximity, both the Alzheimer’s diseases and AN1792 treated groups had the highest levels of post-synaptic colocalization with pTau217 at >100 μm from plaques (**Figure 6**).

**Figure 5:**
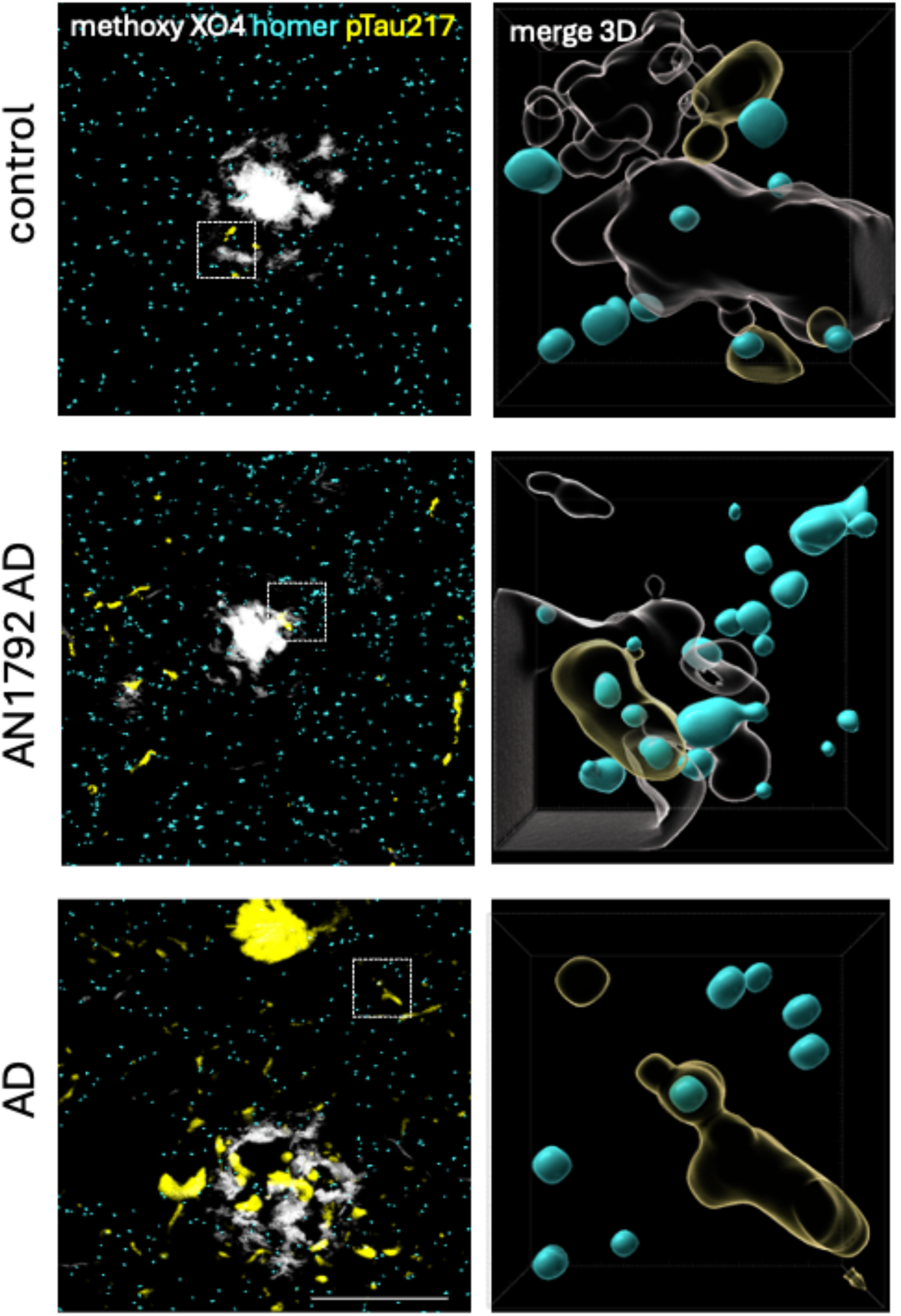
Excitatory post-synapses and pTau217 staining. Methoxy XO4 (white) was used to stain plaque fibrils, Homer 1 (cyan) was used to stain excitatory post-synapses and tau phosphorylated at amino acid 217 (pTau217) was stained (yellow). Images on the left are projections of segmented image stacks. Insets on the right show 3D reconstructions demonstrating homer 1 and pTau217 colocalisation. Scale bar 20 μm, insets 10×10 μm.

**Figure 6:**
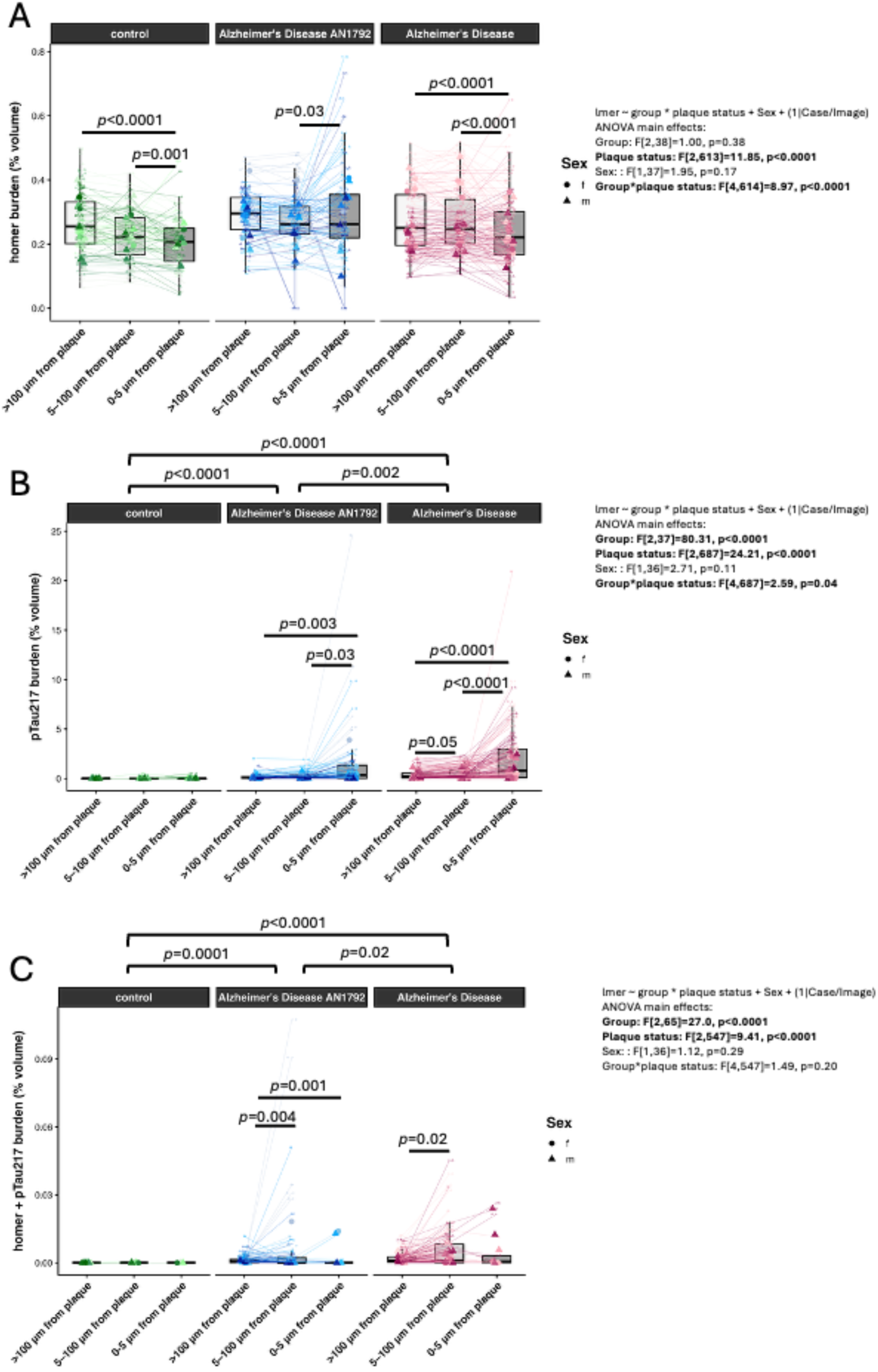
Plaque-associated post-synapse loss and pTau 217 pathology are reduced by AN1792 treatment. Quantification of homer staining reveals plaque-associated loss of post-synapses in control and Alzheimer’s disease but an increase in homer burden within 5 μm of plaques in AN1792 treated Alzheimer’s cases (**A**). Both Alzheimer’s and AN1792 treated Alzheimer’s cases have plaque-associated increases in pTau217 burden, but AN1792 treated cases have significantly lower pTau217 burden (**B**). Colocalisation between pTau217 and homer was highest in the region 5-100 μm from plaques in Alzheimer’s and AN1792 treated Alzheimer’s cases and was significantly higher in both Alzheimer’s and AN1792 treated cases than controls (**C**).

## Discussion

Our results show that even up to 14 years after treatment, vaccination against Aβ is associated with persistent reduction in plaque-associated synapse loss and phospho-tau accumulation. Previous work on these cases demonstrated that AN1792 immunisation was associated with reduced neurite curvature and exacerbated neuron loss, suggesting that vaccination accelerates loss of damaged degenerating neurons thus creating a healthier neuronal environment ^23^. A hypothesis consistent with this idea of immunotherapy creating healthier neuronal networks is that synaptic loss could be reduced or synaptogenesis induced by lowering synaptotoxic Aβ levels. Work in mice demonstrated that both active and passive immunotherapy can prevent amyloid-induced synapse loss ^24^. We also observed that passive immunotherapy can increase formation of new synapses in mice demonstrating synaptic resilience after removal of Aβ ^25^. A previous neuropathological study after aducanumab Aβ antibody immunotherapy reported no change in synapse density in treated patients ^7^; however this study did not look at plaque-associated synapse loss.

Our current data indicate that immunization prevents further pre- and post-synapse loss around existing plaques and/or induces compensatory synaptic plasticity. We speculate that this is due to an ongoing anti- Aβ immune response that promotes microglial clearance of Aβ from the brain thus preventing further synapse loss and allowing the brain’s natural plasticity mechanisms to compensate for neuronal loss by forming new synapses. It is possible that immunization also prevented synapse loss around newly formed plaques after immunisation, however the observation that the AN1972 immunised patients still had detectable antibodies some years after treatment makes new plaque formation unlikely ^26^.

Previous work has shown reduced phospho-tau levels in neurites and reduced astrogliosis with AN1792 vaccination in this cohort ^9,27^. Here we expanded this data to include measures of both AT8 and pTau217 in the immediate vicinity of plaques. We observe that vaccination reduced phospho-tau staining both near and far from plaques. pTau217 is an important form of tau to study in the brain as it is the most widely used fluid biomarker for Alzheimer’s disease, and its levels in blood correlate with plaque accumulation in brain ^28–30^, likely reflecting accumulation of pTau217 in dystrophic neurites around plaques. We further examined astrogliosis and localisation of pathological tau within astrocytes around plaques for the first time after immunization. Immunization was associated with reduction of GFAP burden back to near control levels both near and far from plaques and with a loss of plaque-associated increases in astrocytes containing tau.

It is of interest to note that in our control cases, although there are fewer plaques, those present still induce local synapse loss despite very low levels of phospho-tau and astrogliosis around plaques. This is likely due to direct synaptotoxicity of Aβ and the accumulation of oligomeric Aβ in synapses in a halo around plaques even in control subjects which we have observed previously ^18^. This is consistent with the lack of plaque-associated synapses loss in AN1792 immunised cases as we predict the presence of anti-Aβ antibodies induced by the treatment keeps levels of soluble oligomeric Aβ low as well as clearing amyloid plaques.

A strength of this study is the use of well-characterised human *post-mortem* brain tissue collected up to 14 years after anti-amyloid immunotherapy, enabling the use of high-resolution imaging to examine the long-term effects of amyloid removal on synapses, tau pathology and astrogliosis in the human Alzheimer’s disease brain. However, although our analyses of human *post-mortem* tissue provide direct insight into the long-term effects of anti-amyloid immunotherapy on Alzheimer’s disease pathology, their observational nature precludes establishing the causal mechanisms responsible for preserving synapses and reducing tau pathology. Future work in model systems is needed to fully understand synaptic resilience with amyloid immunotherapies and how this is related to tau pathology and gliosis.

These results are important as they demonstrate long-term protection of synapses with immunotherapy in Alzheimer’s disease. Approved passive immunotherapies lecanemab and donanemab and emerging anti-amyloid therapies in clinical trials also act through lowering amyloid pathology, lending hope to these treatments also promoting synapse resilience as we observe in this active immunisation cohort.

## Data Availability

All data produced in the present study are available upon reasonable request to the authors

## Declarations

MT is an employee of Scottish Brain Sciences. TSJ has received payments for consulting, scientific talks, or collaborative research over the past 10 years from AbbVie, Sanofi, Merck, Boehringer Ingelheim, Eisai, Scottish Brain Sciences, Jay Therapeutics, Cognition Therapeutics, Ono, Bristol Myers Squibb. She is also Charity trustee for the British Neuroscience Association and serves as scientific advisor to several charities and non-profit institutions. DB has been a consultant/advisor for Elan Pharmaceuticals and Biogen and serves as the Editor-in-Chief for Alzheimer’s Research and Therapy. JN has been consultant/advisor in relation to Alzheimer immunotherapy trials for Elan Pharmaceuticals, GlaxoSmithKline, Novartis, Roche, Jansen, Pfizer, Biogen and Eisai.

## Acknowledgements

This work was supported by Alzheimer’s Research UK PhD studentship ARUK-PhD2023-002, the UK Dementia Research Institute (award number UK DRI-4204, to TS-J), through UK DRI Ltd, principally funded by the UK Medical Research Council, the Wellcome Trust Edinburgh Clinical Academic Track for Veterinary Surgeons (to RIM, 225442/Z/22/Z), and the confocal microscope was funded by Alzheimer’s Research UK (ARUK-EG2016A-6). We would like to thank brain tissue donors and their families for providing tissue donations, without which this work would not be possible. We gratefully acknowledge the British Neuroscience Association, Edinburgh Neuroscience and the FENS-Kavli Network of Excellence for facilitating collaborations leading to this work. We gratefully acknowledge the contributions of the Edinburgh Brain and Tissue Bank and Alzheimer’s Scotland Dementia Research Centre for coordinating post-mortem brain tissue donations. The immunised AD samples were obtained from Neuropathology, Cellular Pathology, University Hospital Southampton NHS Trust as part of BRAIN UK, which is supported by Brain Tumour Research and has been established with the support of the British Neuropathological Society and the Medical Research Council.

## Author contributions

Conceptualization (TSJ, DB); Investigation (ES, TSJ); Methodology (MT); Project Administration (JT); Formal Analysis (TSJ); Wrote the first draft of the paper (TSJ); Edited the paper (TSJ, DB, JN, ES) Provided samples with associated data and interpretation (CS, JN, DB).

